# High Incidence of Venous Thrombosis in Patients with Moderate to Severe COVID-19

**DOI:** 10.1101/2020.06.12.20129536

**Authors:** Oleg B. Kerbikov, Pavel Yu. Orekhov, Ekaterina N. Borskaya, Natalia S. Nosenko

**Affiliations:** Federal State Clinical Research Hospital FMBA of Russia; Burnasyan Federal Medical Biophysical Center FMBA of Russia

**Keywords:** COVID-19, Deep vein thrombosis, Thromboprophylaxis

## Abstract

COVID-19 predisposes to venous thromboembolism and there are multiple data regarding high incidence of venous thrombosis in critical COVID-19 patients, however reports on this complication in less severe patients are not widely available.

The aim of this study was to investigate the incidence of deep-vein thrombosis (DVT) in patients with moderate to severe COVID-19 and to assess the prevalence of DVT with lung computerized tomography (lung CT) exams, clinical information and lab data. This study examined 75 consecutive patients with moderate to severe COVID-19, with specific exclusions.

**METHODS:** Almost all patients (pts) admitted to our hospital in the first half of May underwent comprehensive vein ultrasonography. 75 pts (aged 27-92 y, median – 63 y, 36 males and 39 females) with moderate to severe COVID-19 were included in our study.

**RESULTS:** Spontaneous echo contrast (decreased blood velocity and blood stasis) was detected in common femoral veins in 53 pts (70.7%). DVT was found in 15 pts (20%). The vast majority of those with DVT (13 pts, 86.7%) had thrombi only in calf veins and ileofemoral thrombosis was detected in 2 pts with DVT (13.3%). There was no significant observed difference between DVT and non-DVT patients with respect to age, underlying diseases, lung CT scores and SpaO2 at admission. There was also no significant observed difference between DVT and non-DVT patients with respect to both “time from symptoms onset to admission” and with respect to the majority of lab data.

However, a significant difference was observed in D-dimer level (1.87 ± 1.62 vs 0.51 ± 0,4 mcg/mL p<0.0001) and C-reactive protein (116.9 ± 83,6 and 65.1 ± 64.98 mg/L, p = 0.014) for patients with DVT and patients without DVT respectably (Receiver operating characteristics (ROC) curve analysis revealed that the level of D-dimer ≥ 0.69 mcg/mL is the predictor of DVT with a sensitivity of 76.9%, a specificity of 77.6%, p < 0.001 (AUC area under curve = 0.7944). Logistic regression confirmed that D-dimer is an independent predictor of DVT and patients with D-dimer ≥ 0.69 mcg/mL have odds ratio (OR) of developing DVT = 5.1 (confidence interval [CI] 1.9 - 13.5)).

**CONCLUSION:** Patients with moderate to severe COVID-19 show high incidence of DVT, indicating that moderate to severe COVID-19 patients may require an early administration of anticoagulation therapy as part of their treatment. Such therapy may be continued after hospital discharge. Based on these findings, these patients may also require a follow-up with vein ultrasonography after recovery to rule out DVT.

## INTRODUCTION

Coronavirus disease 2019 (COVID-19) is actively spreading globally. While in many cases, COVID-19 presents itself as pneumonia [1], it also affects different organs and results in multiple systemic manifestations including cardiovascular [2], neurological [3] and others [4]. In many COVID-19 patients, abnormal coagulation variables have been demonstrated with high CRP, leukopenia, lymphocytopenia, mild thrombocytopenia, prolonged PT, high D-dimers, and high fibrinogen levels [5, 6].

Several studies revealed high incidence of VTE (venous thromboembolic complications) in patients with severe and critical COVID-19. Wichmann et al [7] found venous thrombosis in 7 out of 12 consecutive autopsies of patients with COVID-19, while Nahum J, etc. found DVT (deep vein thrombosis) in 65% of patients (22 out of 34) admitted to ICU (ultrasonography exam was performed at admission) [8].

Currently, the overwhelming majority of these reports are dedicated to critical COVID-19 patients and there is lack of data regarding patients with moderate to severe (but not critical) COVID-19.The aim of this study was to evaluate incidence of DVT in patients with moderate to severe COVID-19 and to assess the prevalence of DVT with clinical findings and laboratory data.

## METHODS

Almost all patients admitted to Federal State Clinical Research Hospital FMBA of Russia with COVID-19 in the first half of May 2020 underwent venous ultrasonography using the full venous duplex ultrasonography (VDU) protocol which includes scanning of the deep venous system from the iliac veins to the calf veins bilaterally as well as peripheral veins using gray-scale (B-mode) and color flow, spectral analysis, and maneuvers of compression and augmentation, as outlined in standards published by the Society for Vascular Ultrasound (SVU) [9] and Intersocietal Accreditation Commission (IAC) [10] with the exception of Valsalva maneuver. All studies were performed by credentialed vascular technologist (ARDMS RVT). All examinations were carried out within 7 days following hospital admission.

Critical COVID-19 patients who required admission to ICU, patients with more than 75% of lungs affected (as evidenced by lung CT exam), patients with a known history of DVT and patients with recent leg and hip trauma were excluded from analysis. A total of 75 consecutive patients were included for further evaluation, after exclusions.

Statistical analyses were conducted in NCSS 2020 (NCSS, LLC) and Excel 2007 (Microsoft Corp). Statistical significance was set at *p*< .05, and all tests were 2-tailed.

Patient characteristics are outlined in Table 1.

**Table 1.**
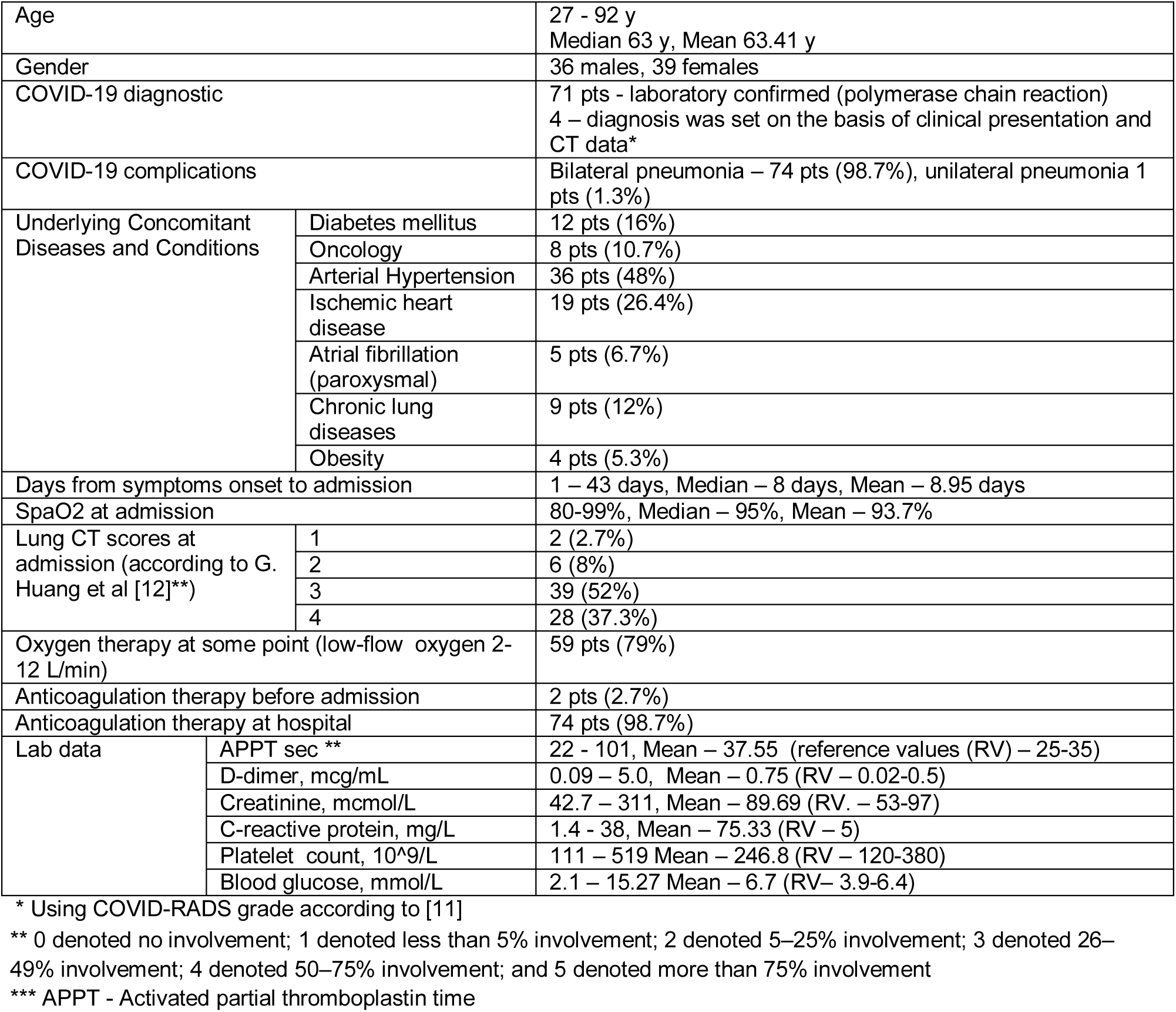
Patient characteristics (75 Patients)

## RESULTS

Vein ultrasonography revealed spontaneous echo contrast in common femoral veins in 53 pts (70.7%) which indicated blood stasis and markedly decreased venous flow velocity in majority of patients. Vein thrombosis was found in 16 patients - 1 pts had superficial thrombosis and 15 pts (20%) had DVT. 4 pts had bilateral DVT and 11 unilateral. Characteristics of thrombi are described in table 2.

**Table 2.** Patients with vein thrombosis. Affected veins and characteristics of thrombi

|  | Number of Affected veins | Pts |
| --- | --- | --- |
| External Iliac vein | 2 | 2 |
| Common femoral vein | 2 |  |
| Superficial femoral vein | 2 |  |
| Popliteal vein | 1 | 1 |
| Sural vein | 4 | 15 |
| Posterior tibial veins | 7 |  |
| Peroneal veins | 10 |  |
| Venous sinuses | 5 |  |
| Superficial veins | 4 | 4 |
| Occlusive thrombosis (DVT) | 21 |  |
| Non-occlusive thrombosis (DVT) | 12 |  |
| Floating thrombus | 2 (thrombi in external iliac veins) |  |
| Bilateral DVT | 4 pts |  |
| Unilateral DVT | 11 pts |  |
| Clinical manifestations (pain, swelling, red or darkened skin etc) | 1 pts |  |

A majority of patients were found to have thrombi in calf veins only (13 pts, 86.7%) and 2 pts (13.3%) have ileofemoral thrombosis (one pt has also popliteal vein involvement, and both pts have calf vein thrombosis). These same 2 pts have floating thrombi with a high risk of pulmonary embolism. Most of the thrombi were occlusive thrombi in the peroneal, posterior tibial veins and sinuses (fig 1).

Patients with DVT (Group 1) were somewhat older than those without thrombi (Group 2) and they a had longer time between symptoms onset and admission to hospital (Table 3). While noted, however, those differences were not significant. There was no significant difference in the share of patients with underlying concomitant diseases and conditions (diabetes mellitus, oncology, ischemic heart disease etc.) between these groups. There was also no difference in SpaO2 values and lung lesions (measured using lung CT scores according to [12]). However, patients with DVT had significantly higher D-dimer and C-reactive protein levels (Table 3). In accordance with recommendations, almost all patients received anticoagulant prophylaxis at hospital admission, however only 2 pts had received such therapy before admission.

**Table 3.** Comparison between patients with DVT (Group 1) and those without thrombi in the veins of the deep lower extremities (Group 2).

|  |  | Patients with DVT (Group 1)<br>15 pts | Patients without DVT (Group 2)<br>60 pts | p | Std. Dev. *<br>(Group 1) | Std. Dev.<br>(Group 2) |
| --- | --- | --- | --- | --- | --- | --- |
| Age (years) |  | 62 | 69.1 | 0.087 | 15.21 | 14.03 |
| Underlying Concomitant Diseases and Conditions | Diabetes mellitus (pts) | 2 (13.3%) | 10 (16.7%) | 0.753** |  |  |
|  | Oncology (pts) | 1 (6.7%) | 7 (11.7%) | 0.575** |  |  |
|  | Arterial Hypertension (pts) | 10 (66.7%) | 26 (43.3%) | 0.106** |  |  |
|  | Ischemic heart disease (pts) | 6 (40%) | 13 (21.7%) | 0.144** |  |  |
|  | Atrial fibrillation (paroxysmal) (pts) | 2 (13.3%) | 3 (5%) | 0.259** |  |  |
|  | Chronic lung diseases (pts) | 1 (6.7%) | 8 (13.3%) | 0.477** |  |  |
| Time from symptoms onset to admission (days) |  | 11.2 | 8.4 | 0.153 | 9.71 | 5.59 |
| SpaO2 at admission (%) |  | 94 | 92.6 | 0.192 | 3.9 | 3.5 |
| Lung CT scores at admission |  | 3.4 | 3.2 | 0.335 | 0.63 | 0.73 |
| APPT sec *** |  | 40.5 | 36.9 | 0.34 | 11.46 | 12.6 |
| D-dimer, mcg/mL *** |  | 1.87 | 0.51 | **<0.0001** | 1.62 | 0.4 |
| Creatinine, mcmol/L *** |  | 83.96 | 90.9 | 0.643 | 60.36 | 43.8 |
| C-reactive protein, mg/L *** |  | 116.9 | 65.1 | **0.014** | 83.6 | 64.98 |
| Platelet count, 10<sup>9</sup>/L *** |  | 272.7 | 239.7 | 0.203 | 85.0 | 85.3 |
| Blood glucose, mmol/L *** |  | 6.0 | 6.9 | 0.185 | 1.3 | 2.3 |

\* Std. Dev. – standard deviation (where applicable)
\*\* Calculated using Chi-square test
\*\*\* For reference values see Table 2.

Receiver operating characteristics (ROC) curve analysis revealed that the level of D-dimer ≥ 0.69 mcg/mL is the predictor of DVT with a sensitivity of 76.9%, a specificity of 77.6%, p < 0.001 (AUC area under curve = 0,7944). Using a C-reactive protein as predictor resulted in much less accuracy (AUC = 0.68 sensitivity 66.7%, specificity – 63.6%, C-reactive protein level ≥70.1 mg/L) (Fig 2).

**Fig 2.**
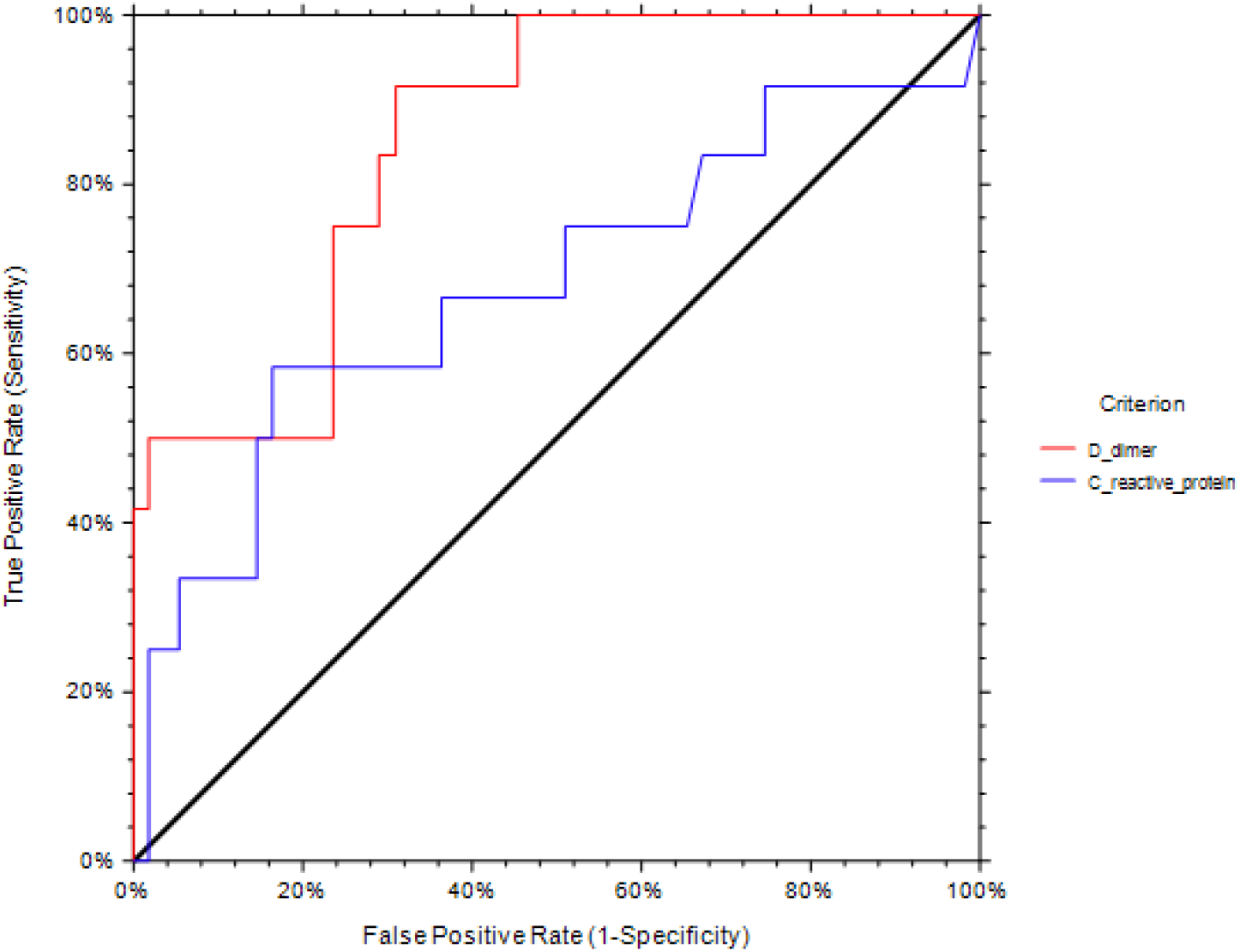
ROC curve of DVT

Logistic regression confirmed that D-dimer level is an independent predictor of DVT controlling gender, age, Lung CT score, SpaO2 at admission, Platelet count, APPT, Creatinine and C-reactive protein (Chi Square= 18.3038; df=8; p= 0.0191). Patients with D-dimer ≥ 0.69 mcg/mL have an enhanced risk of developing DVT odds ratio (OR) = 5.1 (confidence interval CI – 1.9-13.5).

## DISCUSSION

Frequent incidence of venous thrombosis (up to 27-65% of all patients) was reported in critical COVID-19 patients [8, 13]. In contrast there is not enough data about frequency of DVT in patients with less severe form of disease. In our study of moderate to severe consecutive patents we found 20% incidence of DVT in patients with moderate to severe COVID-19 and most of the patients have decreased venous velocity and blood stasis. With respect to thrombi, the majority are occlusive thrombi in calf veins and while these pose a somewhat reduced risk of pulmonary embolism, they are very difficult to treat. Despite intensive anticoagulant treatment with low molecular weight heparins (like nadroparin) there were limited results and all thrombi persisted, which is consistent with [8]. Two of patients had ileofemoral thromobosis with an accompanied high risk of pulmonary embolism, which was averted thanks to early detection. Also, in two of patients with DVT, we detected thrombi in upper extremities veins which highlights the possibility of widespread thrombi formations in patients even with only moderate COVID-19. The majority, 65 pts (86.7%), were discharged from hospital and those with DVT continue anticoagulant treatment. The D-dimer level and C-reactive protein level were increased in the majority of COVID-19 patients which is consistent with published data [5, 6]. However, this increase was more pronounced in patients with DVT and, based on our observations, we proposed a D-dimer level threshold of 0.69 mcg/mL as a level representing significantly increased risk of DVT however this requires further study. The majority of patients 65 pts (86.7%) were discharged from the hospital and those with DVT continue anticoagulant treatment. 10 patients (13.3%) continue treatment in hospital. Perhaps it is advisable to start anticoagulation therapy as early as possible in patients with moderate to severe COVID-19 and continue it after discharge from hospital. Also, it perhaps makes sense to follow these patients and, at some point in the future, to perform venous ultrasonography to rule out DVT.

## Data Availability

All data will be available upon request

## Author Contributions

Drs. Oleg Kerbikov Pavel Orekhov and Ekaterina Borskaya. Drs had full access to all of the data in the study and take responsibility for the integrity of the data and the accuracy of the data analysis.

*Concept and design:* Oleg Kerbikov Pavel Orekhov Natalia Nosenko

*Acquisition, analysis, or interpretation of data:* Oleg Kerbikov Ekaterina Borskaya Pavel Orekhov

*Drafting of the manuscript:* Oleg Kerbikov, Ekaterina Borskaya

## Notes

### Competing Interest Statement

The authors have declared no competing interest.

### Funding Statement

No funding

### Author Declarations

Local Hospital IRB

